# Ethnicity is not biology: retinal pigment score to evaluate biological variability from ophthalmic imaging using machine learning

**DOI:** 10.1101/2023.06.28.23291873

**Authors:** Anand E Rajesh, Abraham Olvera-Barrios, Alasdair N. Warwick, Yue Wu, Kelsey V. Stuart, Mahantesh Biradar, Chuin Ying Ung, Anthony P. Khawaja, Robert Luben, Paul J. Foster, Cecilia S. Lee, Adnan Tufail, Aaron Y. Lee, Catherine Egan, EPIC Norfolk, UK Biobank Eye and Vision Consortium

**Affiliations:** Department of Ophthalmology, University of Washington, Seattle, WA, USA; The Roger and Angie Karalis Johnson Retina Center, Seattle, WA, USA; NIHR Biomedical Research Centre, Moorfields Eye Hospital NHS Foundation Trust & University College London Institute of Ophthalmology, London, UK; University of Cambridge, Cambridge, UK; Guy’s and St Thomas’ NHS Foundation Trust; MRC Epidemiology Unit, University of Cambridge, Cambridge, UK

## Abstract

**Background:** Few metrics exist to describe phenotypic diversity within ophthalmic imaging datasets, with researchers often using ethnicity as an inappropriate marker for biological variability.

**Methods:** We derived a continuous, measured metric, the retinal pigment score (RPS), that quantifies the degree of pigmentation from a colour fundus photograph of the eye. RPS was validated using two large epidemiological studies with demographic and genetic data (UK Biobank and EPIC-Norfolk Study).

**Findings:** A genome-wide association study (GWAS) of RPS from UK Biobank identified 20 loci with known associations with skin, iris and hair pigmentation, of which 8 were replicated in the EPIC-Norfolk cohort. There was a strong association between RPS and ethnicity, however, there was substantial overlap between each ethnicity and the respective distributions of RPS scores.

**Interpretation:** RPS serves to decouple traditional demographic variables, such as ethnicity, from clinical imaging characteristics. RPS may serve as a useful metric to quantify the diversity of the training, validation, and testing datasets used in the development of AI algorithms to ensure adequate inclusion and explainability of the model performance, critical in evaluating all currently deployed AI models. The code to derive RPS is publicly available at: https://github.com/uw-biomedical-ml/retinal-pigmentation-score.

**Funding:** The authors did not receive support from any organisation for the submitted work.

**Research in context:** *Evidence before this study:* Vision loss due to retinal disease is a global problem as populations age and diabetes becomes increasingly prevalent. AI algorithms developed for efficient diagnosis of diabetic retinopathy and age-related macular degeneration rely on large imaging datasets collected from clinical practice. A substantial proportion (more than 80%) of publicly available retinal imaging datasets lack data on participant demographic characteristics. Some ethnic groups are noticeably underrepresented in medical research. Previous findings in dermatology suggest that AI algorithms can show reduced performance on darker skin tones. Similar biases may exist in retinal imaging, where retinal colour has been shown to affect disease detection.

*Added value of this study:* We introduce the Retinal Pigment Score (RPS), a measure of retinal pigmentation from digital fundus photographs. This score showed strong, reproducible associations with genetic variants related to skin, eye, and hair colour. Additionally, we identify three genetic loci potentially unique to retinal pigmentation, which warrant further investigation. The RPS provides an accurate and objective metric to describe the biological variability of the retina directly derived from an image.

*Implications of all the available evidence:* The RPS method represents a valuable metric with importance to harness the detailed information of ophthalmic fundus imaging. Its application implies potential benefits, such as improved accuracy and inclusivity, over human-created sociodemographic classifications used in dataset compilation and in the processes of developing and validating models. The RPS could decouple the distinct social and political categorical constructs of race and ethnicity from image analysis. It is poised to both accurately describe the diversity of a population study dataset or an algorithm training dataset, and for investigate algorithmic bias by assessing outcomes. Further work is needed to characterise RPS across different populations, considering individual ocular factors and different camera types. The development of standard reporting practices using RPS for studies employing colour fundus photography is also critical.

## Introduction

Retinal diseases are a significant global cause of vision loss, but not all populations are affected equally. In 2020, there were estimated to be 103·1 million adults worldwide with diabetic retinopathy (DR) and 196 million people with age-related macular degeneration (AMD).[1] Studies have found DR prevalence is highest in Africa (35·9%), then North America and the Caribbean (33·3%). In contrast, AMD has a significantly higher prevalence in people of European than in those of Asian or African ancestry.[2,3] In response to the overwhelming global burden of disease, many artificial intelligence (AI) algorithms have been developed to enable more efficient care delivery. These AI algorithms have been widely published, and several are already in clinical practice for automated diagnoses for DR, AMD, and glaucoma.[4–7]

The success of AI algorithms in ophthalmology is partly due to the availability of large imaging datasets that have been collected from routine clinical practice.[8] Less than 20% of publicly available retinal imaging datasets contain patient characteristics such as age, sex, or ethnicity.[8] Studies that compare model performance across different populations are severely limited.[4,9,10]

Bias in image-based AI algorithms is often worse among people with a greater degree of skin pigmentation, for instance, in skin cancer classification, facial recognition, and object detection.[11–13] Studies use a multi-step categorical pigmentation scale, or even a binary, “light” vs “dark” skin tone classification, and often with humans labelling images of other humans. These categories are then used to estimate relative performance of algorithms within subcategories of pigmentation.

In the eye, melanin is present in the uvea (iris, retina, and choroid) and is responsible for blue or brown iris colour as well as retinal pigmentation.[14,15] We aimed to develop a continuous scale, the Retinal Pigment Score (RPS), to quantify the background pigmentation of retinal colour fundus photographs. We then sought to validate the RPS by comparing with self-reported ethnicity and performing both genome-wide (GWAS) and phenome-wide (PheWAS) association studies. Mechanistic insight was attained through gene priorisation and functional annotation whilst causal associations with clinically relevant outcomes were tested using mendelian randomisation (MR). Finally, we validated our results with a replication GWAS study in a separate cohort.

## Methods

### Ethics

We analysed data from UK Biobank participants who as part of their examinations underwent enhanced ophthalmic review. Ethics approval was obtained by the Northwest Multi-centre Research Ethics Committee (REC reference number 06/MRE08/65; approved project number 28541).

The European Prospective Investigation into Cancer and Nutrition-Norfolk (EPIC-Norfolk) Eye Study was carried out following the principles of the Declaration of Helsinki and the Research Governance Framework for Health and Social Care and was approved by the Norfolk Local Research Ethics Committee (identifier: 05/Q0101/191) and the East Norfolk and Waveney National Health Service Research Governance Committee (identifier: 2005EC07L).

### Study population

The UK Biobank is a national research resource aiming to improve prevention, diagnosis, and treatment of a wide range of diseases. More than 500,000 people aged 37-73 were recruited at 22 study assessment centres across the UK between January-2006 and October-2010. The EPIC-Norfolk Eye Study comprises 8,623 participants aged 40 to 79 years from Norfolk, England.[16] Participants were recruited from 35 participating general practices. Baseline examinations were carried out between 1993 and 1997.[16]

### Ophthalmic assessment

In the UK Biobank, more than 133,000 participants underwent an enhanced ophthalmic assessment between 2009 and 2010 at 6 assessment centres, including ophthalmic imaging.[17] The right eye was imaged first. Single-field colour fundus photographs (45° field-of-view, centred to include both optic disc and macula) and macular OCT scans were captured using a digital Topcon-1000 integrated ophthalmic camera (Topcon 3D OCT1000 Mark II, Topcon Corp., Tokyo, Japan). Ophthalmic examination for 8,623 EPIC-Norfolk participants was performed between 2004 and 2011. Fundus photography was acquired using a TRC-NW6S non-mydriatic retinal camera and IMAGEnet Telemedicine System (Topcon Corporation, Tokyo, Japan) with a 10 megapixel Nikon D80 camera (Nikon corporation, Tokyo, Japan).[16]

### Retinal Pigment Score

Figure 1 represents a schematic of the pipeline. Each fundus image was run through the Automorph pipeline[18] to create a segmentation mask of the retinal vasculature and optic disc. In summary, each image is pre-processed then passed through an image quality classifier. Images of insufficient quality are excluded from the subsequent segmentation steps. We modified the Automorph code by changing the file system organisation between input and output nodes, adding in error handling, and reducing output file size.

**Figure 1.**
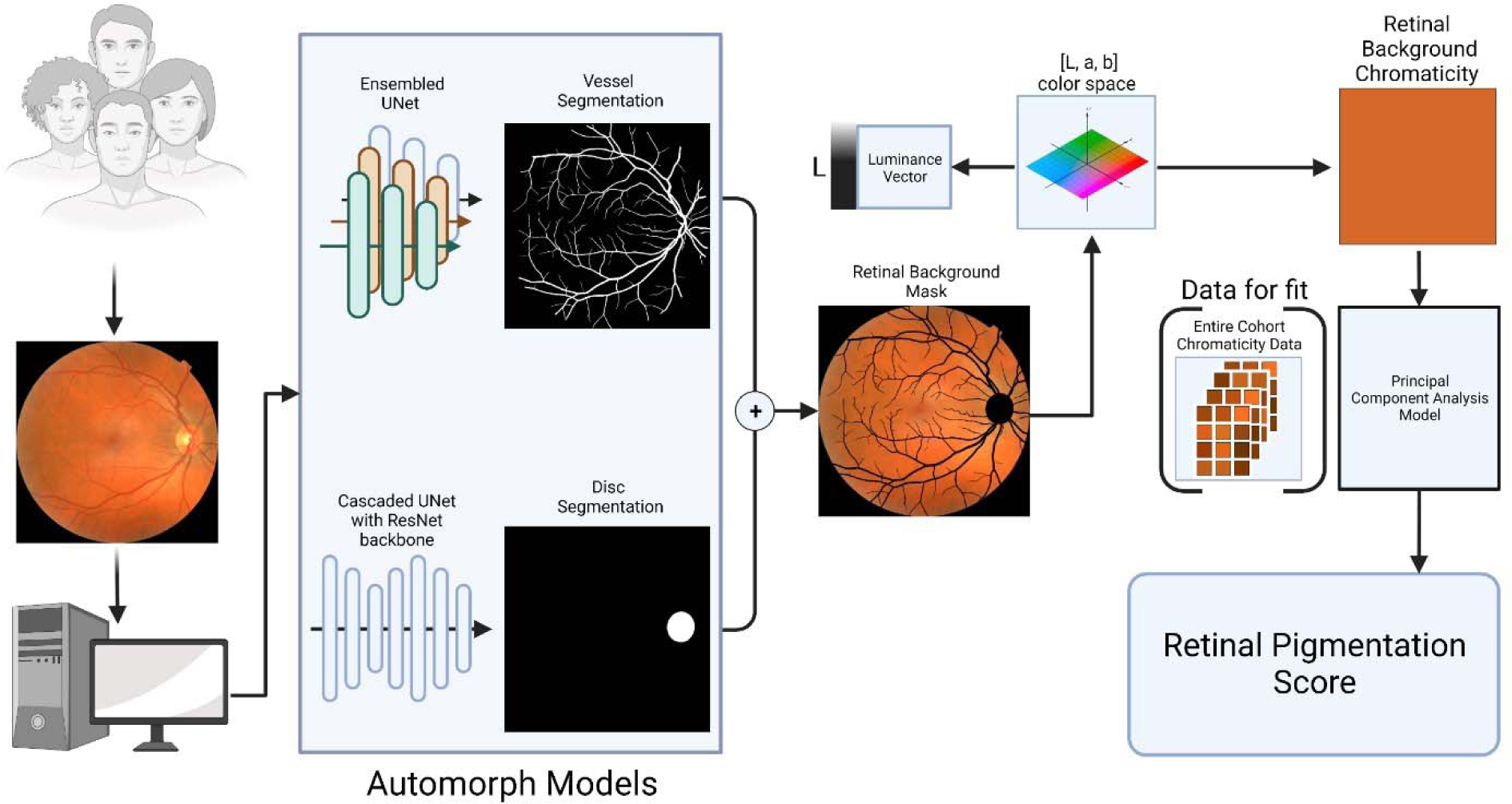
Schematic showing the method to generate the retinal pigmentation score (RPS) from a colour fundus image. Input images are fed into the deep learning algorithm to generate segmentation masks. These are added together to make a retinal background mask, which is then transformed into L,a,b colorspace. The chromaticity vectors are then extracted and transformed by a principal component analysis model to create the RPS.

We added the segmentation masks for the disc segmentation and the binary vessel segmentations to make a combined disc/vessel mask. The background was identified by finding all pixels that were at or below the 0·5 percentile of the distribution of all grayscale pixels from the input image. The background mask was added to the combined vessel/disc mask. This mask was then successively dilated using a 2-dimensional binary structuring kernel with connectivity of 2. The number of dilation iterations was 4 multiplied by pixel width of the image divided by 600 rounded to the nearest integer, which was derived empirically. All pixels not contained in dilated masks of the background/disc/segmentation mask were used to create a new retinal background mask.

From the retinal background mask, the median RGB pixel value was converted into CIE-LAB colour space which is a colour space designed to have luminance (L vector) stored in a separate vector from chromaticity (a,b vectors).[19] To reduce the effect of illumination, we only used the a,b coordinates from the CIELAB space and ignored the L vector. To transform the two dimensional a,b chromaticity vectors for each eye, we used a principal component analysis (PCA) model to perform dimensionality reduction. For each dataset, a two component PCA model was fitted to the median a,b value of the retinal background for all images in the dataset. Then, each eye’s median a,b value was transformed with the PCA model along the eigenvector with the greatest eigenvalue. This new transformed vector was stored as the 1-dimensional RPS vector. The image analysis to derive the RPS was performed with Python, version 3·8[20] and PyTorch, version 1·7.0.[21] The code to derive RPS is publicly available at: https://github.com/uw-biomedical-ml/retinal-pigmentation-score.

### Genome-Wide Association Study

GWAS was performed to assess potential genetic associations with mean RPS (average score between right and left eyes per participant). A beta coefficient of 1 therefore corresponds to a 1 standard deviation increase in standardised mean RPS. Analyses were conducted using a generalised linear mixed model, adjusting for age, sex, and the first ten principle components. The initial discovery GWAS analysis was performed in the UK Biobank cohort. Lead variants reaching genome-wide significance (p<5×10^-8^) were re-evaluated in a replication GWAS analysis, conducted in the EPIC-Norfolk cohort.

Lead variants were furthermore investigated for previously identified associations with hair, skin and eye colour, by manually referring to the Open Targets Genetics[22,23] and PhenoScanner[24,25] resources, and listed in table 1. Further details for GWAS analysis are presented as supplements (supplementary methods).

**Table 1.** Genome-wide significant associations with retinal pigment score in the UK Biobank cohort. Variants that met the replication threshold in the EPIC-Norfolk replication GWAS are highlighted in the ‘Replicated’ column. The last 3 columns indicate which variants have previously been shown to be associated with hair, skin or iris colour.

| Rs identifier | chr:pos<br>[hg19] | EA/OA<br>(EAF) | Beta<br>(95%<br>CI) | P | Nearest gene | Hair colour | Skin<br>colour | Eye<br>colour |
| --- | --- | --- | --- | --- | --- | --- | --- | --- |
| rs6670870 | 1:205155177 | A/T (0.76) | -0.09 (-0.11; -0.08) | 8.7E-36 | *DSTYK* | Yes |  |  |
| rs173273 | 1:212446689 | G/T (0.41) | 0.04 (0.02; 0.05) | 2.9E-08 | *PPP2R5A* | Yes | Yes |  |
| rs762948237 | 3:129178587 | TCTTC/T (0.87) | 0.05 (0.03; 0.07) | 2.3E-08 | *IFT122* |  |  |  |
| rs16891982 | 5:33951693 | C/G (0.02) | 0.52 (0.48; 0.56) | 1.5E-135 | *SLC45A2* | Yes | Yes | Yes |
| rs12203592 | 6:396321 | C/T (0.79) | 0.13 (0.11; 0.14) | 2.4E-59 | *IRF4* | Yes | Yes | Yes |
| rs62425803 | 6:134330249 | G/A (0.81) | 0.05 (0.04; 0.07) | 1.4E-11 | *TCF21* | Yes |  |  |
| rs117756744 | 7:100277212 | G/A (0.98) | 0.18 (0.14; 0.23) | 5.4E-17 | *GNB2* | Yes | Yes |  |
| rs1325117 | 9:12613472 | G/A (0.36) | 0.06 (0.05; 0.07) | 3.0E-19 | *TYRP1;LURAP1L* | Yes | Yes |  |
| rs11023814 | 11:16007053 | C/G (0.43) | 0.04<br>(0.03; 0.06) | 2.2E-12 | SOX6 |  | Yes |  |
| rs150527451 | 11:68817897 | G/A (0.89) | 0.16<br>(0.14; 0.18) | 1.7E-53 | TPCN2 | Yes | Yes |  |
| rs1060435 | 11:68855595 | A/G (0.59) | 0.07<br>(0.05; 0.08) | 1.1E-24 | TPCN2 | Yes | Yes | Yes |
| rs747572 | 11:87885082 | A/G (0.63) | 0.05<br>(0.04; 0.06) | 4.9E-14 | CTSC | Yes | Yes |  |
| rs1126809 | 11:89017961 | G/A (0.7) | 0.07<br>(0.06; 0.09) | 5.0E-27 | TYR | Yes | Yes | Yes |
| rs4762973 | 12:20710145 | A/G (0.75) | 0.06<br>(0.04; 0.07) | 7.8E-15 | PDE3A |  |  |  |
| rs10771034 | 12:23979199 | T/A (0.45) | -0.04 (-0.06; -0.03) | 1.1E-12 | SOX5 | Yes | Yes |  |
| rs766338951 | 13:95169060 | CT/C (0.69) | 0.08<br>(0.06; 0.09) | 4.6E-30 | DCT | Yes | Yes |  |
| rs1800407 | 15:28230318 | C/T (0.91) | 0.11<br>(0.09; 0.13) | 1.8E-23 | OCA2 | Yes | Yes | Yes |
| rs12913832 | 15:28365618 | A/G (0.22) | 0.44<br>(0.43; 0.46) | 0.0E+00 | HERC2 | Yes | Yes | Yes |
| rs7220155 | 17:79606020 | C/T (0.62) | -0.06 (-0.07; -0.05) | 9.2E-22 | TSPAN10 | Yes | Yes |  |
| rs1785433 | 21:44783282 | A/G (0.65) | -0.04 (-0.05; -0.02) | 1.3E-08 | SIK1 |  |  |  |

### Phenome-Wide Association Study

A PheWAS analysis was conducted within the discovery UK Biobank GWAS subcohort using 308 CALIBER codelists drawing on the following diagnostic records: verbal interview responses, linked hospital episode statistics, death register, and primary care records. Read-2, ICD-10 and OPCS-4 clinical codelists were minimally adapted from the CALIBER Portal.[26] The former two coding systems were expanded to Read-3 and ICD-9 equivalents respectively, using the mapping files provided by UK Biobank Resource 592 (https://biobank.ndph.ox.ac.uk/ukb/refer.cgi?id=592).

The PheWAS analysis performed logistic regression to assess potential disease associations with RPS, adjusting for age and sex. All available diagnostic records, both before and after the date of attendance for retinal imaging, were included. Conditions with fewer than 200 cases were excluded. Associations meeting the Bonferroni-corrected p-value threshold (p=0·05/308) were considered phenome-wide significant.

### Mendelian randomisation

Main MR analyses (supplementary methods) were performed using a multiplicative random-effects inverse-variance weighted approach.[27]. To account for invalid instrumental variables and pleiotropy, we conducted sensitivity analyses using three alternative MR methods: weighted median, weighted mode, and MR-Egger.

### Sociodemographic and phenotypic associations of RPS

Linear regression models with mean RPS adjusting for age, sex, self-reported ethnicity (categorised as white, black, Asian, mixed, Chinese, or other), hair colour (categorised as blonde, red, light brown, dark brown, black and other), skin colour (categorised as very fair, fair, light olive, dark olive, brown and black), spherical equivalent, height, quintiles of Townsend deprivation index (TDI) (where a higher quintile implies a greater degree of deprivation), and UK Biobank assessment centre were used to examine associations with RPS. Missing data points were categorised as “Missing” within each variable.

## Results

### Retinal Pigment Score

A total of 135,592 colour fundus photographs (67,982 right eyes, 67,610 left eyes) from 68,504 participants were available for analysis. From these, 74,851 images (40,329 right eyes, 34,388 left eyes) of 44,320 participants (55% female) were gradable by our pipeline and included in the analysis. The cohort characteristics are summarised in supplementary table 1. The median age (IQR) was 56 years (49-63) and 92% (40,704/44,320) of participants self-described their ethnicity as white. The median RPS was −0·82 (−9·89, 10·39). Each ethnic group had images in each quintile of the range of RPS, except for Chinese in the lowest (less pigmented) quintile (figure 2).

**Figure 2.**
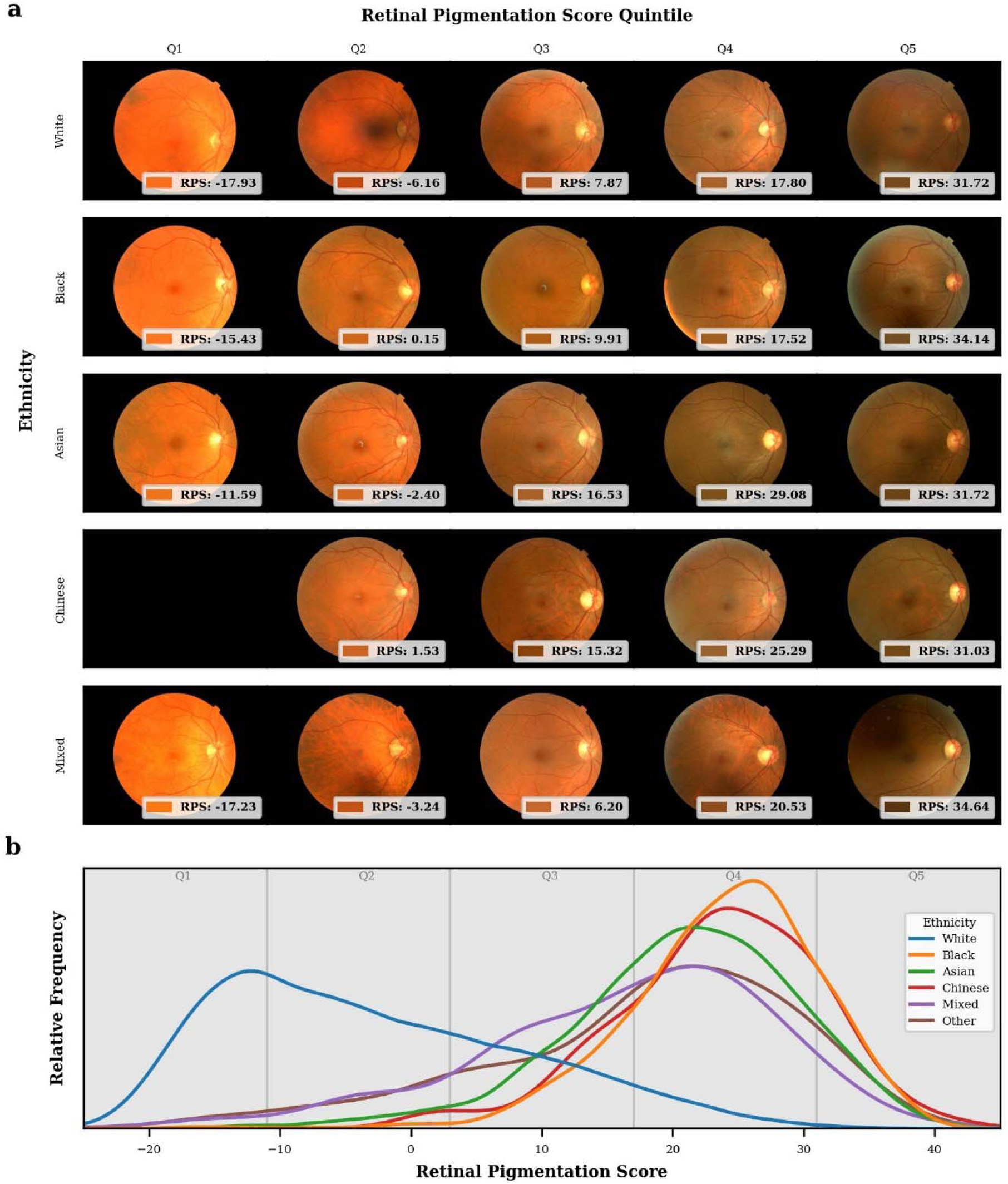
a. Randomly sampled colour fundus photographs from each self-reported ethnicities by quintiles of retinal pigment score (RPS) across the entire distribution of RPS for the UK Biobank cohort and each associated self-reported ethnicity of the participant. The retinal background colour and the RPS is shown at the bottom of each fundus photograph. b. Normalised kernel density estimation plot of the distribution of RPS for all participants grouped by self-reported ethnicity as reported in the UK Biobank. Relative frequencies are normalised so the area under each curve is equal for each ethnicity.

### Associations of RPS with Clinical Variables

Supplementary figure 1 shows the association of RPS with the covariates of interest (deciles of continuous variables) adjusted for age, sex, and UK Biobank centre. Non-white self-described ethnicities were associated with increased RPS when compared to white individuals. A positive graded association was observed with increased skin pigmentation, hair pigmentation, and deprivation. An inverse linear association with RPS was evidenced for height.

Next, the associations were tested with multivariable linear regression adjusting for age, sex, height, self-described ethnicity, self-described hair and skin colour, TDI, refractive status, and UK Biobank assessment centre (supplementary table 2). Every 5-year rise in age was associated with a 0·20 increase in RPS (p 1·3×10^-8^), and every 5cm increase in height conferred a −0·24 change in RPS (p 3·6×10^-8^). A non-linear association was evidenced for refractive status. A higher RPS was observed in people with emmetropia (2·1, 95%CI 1·5-2·6; p 1·1×10^-12^), and hyperopia (1·4, 95%CI 0·84-2; p 1·1×10^-6^) when compared to people with high myopia. The most deprived TDI quintile showed a 0·81 increase in RPS when compared to the least deprived TDI (p-for-linear-trend 3×10^-4^). When compared to very fair skin colour, darker skin tones showed a graded increase in RPS (p-for-linear-trend 2·5×10^-231^). People with black skin colour showed an 11 unit increase in RPS (95%CI 9·4-12; p 5·7×10^-48^) when compared with people of very fair skin colour. Similarly, when compared to people with blonde hair, darker hair colours showed a graded positive association with RPS (p-for-linear-trend 2·2×10^-155^). People with black hair colour showed a 7-point increase in RPS when compared to people with blonde hair colour (p 4·8×10^-122^). There was a strong association of ethnicity with RPS. When compared to white individuals, people self-described as non-white showed a more than 10 unit increase in RPS with Chinese (20, 95%CI 18-21; p 4·6×10^-103^) and black (15, 95%CI 14-16; p 1·6×10^-158^) people showing the largest effect sizes. However, within ethnic groups there was a wide spread of RPS values, overlapping with other groups (figure 2). Sex was not associated with RPS.

### Genome-Wide Association Study

#### Discovery analysis

A GWAS was performed to assess potential associations with standardised mean RPS. The discovery analysis included 37,067 individuals of European ancestry from the UK Biobank cohort. The genomic inflation factor was 1·071, and the linkage disequilibrium score regression intercept was 1·013 with a ratio of 0·09. Conditional analysis identified 20 independent autosomal genomic loci reaching genome-wide significance (p<5×10^-8^), the majority of which have previously been shown to associate with hair, skin and/or iris colour (table 1, figure 3).

**Figure 3.**
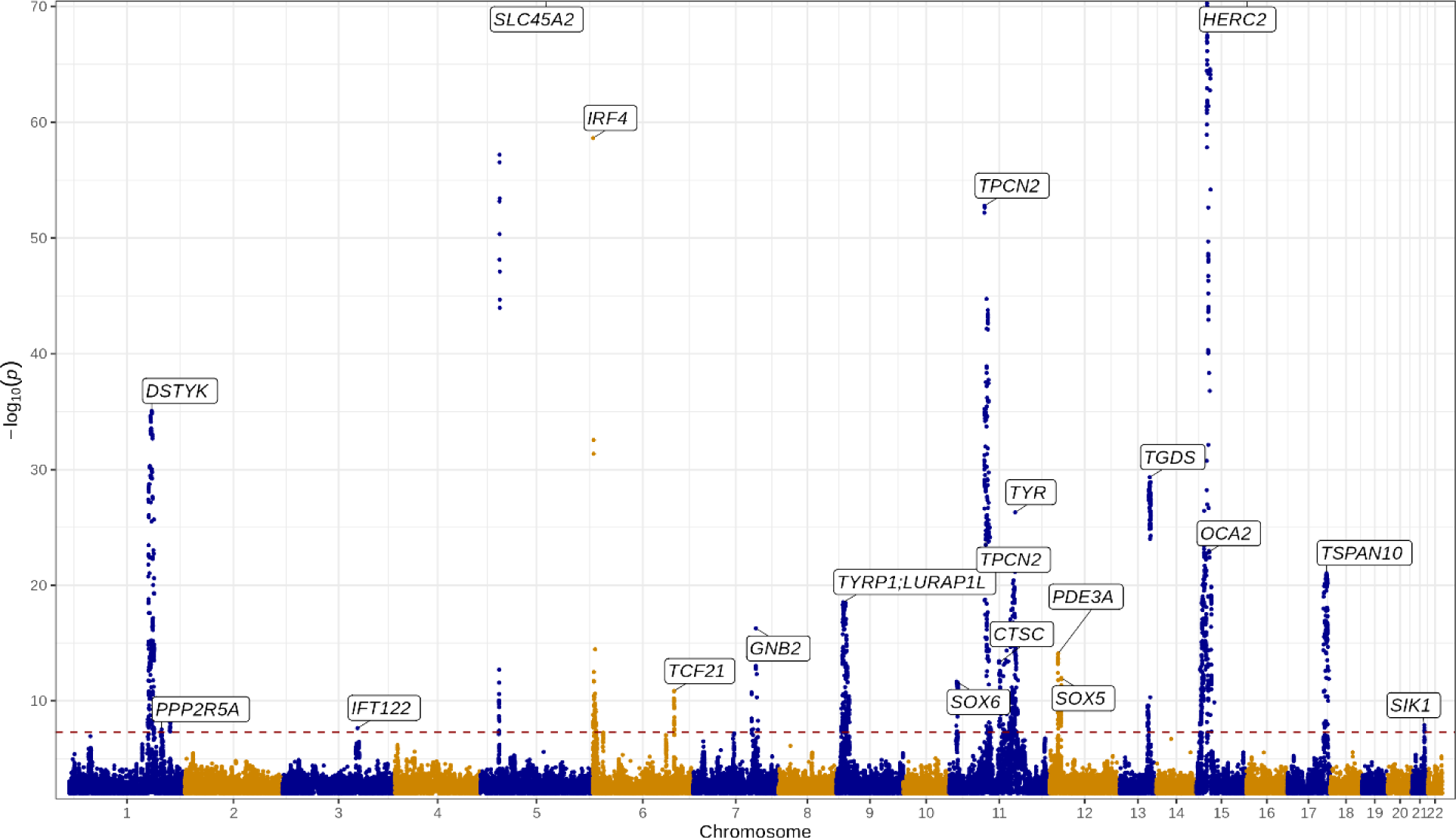
Manhattan plot of GWAS results. Lead variants identified by GCTA-COJO are annotated with the nearest gene. Points are truncated at -log10(p) = 70 for clarity. The dashed red line indicates genome-wide significance (p = 5 x 10^-8^).

Positional and expression quantitative trait locus mapping in retina, skin and dermal fibroblasts were performed to identify candidate causal genes at each independent risk locus. This produced a set of 100 prioritised genes (supplementary table 3) which were then annotated in biological context. A number of these had existing entries in the GWAS Catalog[28], with enrichment for traits including hair, eye and skin colour, as well as various skin malignancies (supplementary figure 2). Enrichment for several Gene Ontology entries was also apparent, especially those related to melanin and pigmentation biological processes (supplementary figure 3).

#### Replication analysis

A replication GWAS was conducted in the independent EPIC-Norfolk Eye Study cohort.[16] The replication analysis included 4,273 individuals of European ancestry. Due to differences in genotyping platforms and imputation methods, three of the lead variants highlighted in the discovery GWAS were either unavailable or did not pass quality control in the replication dataset (rs173273, rs762948237, and rs766338951). Replication was therefore assessed for 17 out of the 20 lead variants.

The direction of effect was concordant for all 17 variants and highly correlated with estimates from the discovery analysis (Pearson’s rho 0·986 [95%CI: 0·961, 0·995]) (figure 4). Of the 17 variants, 15 variants were significant at p<0·05, 8 remained significant after adjusting for multiple testing (p<0·05/17), and 2 achieved genome-wide significance (supplementary table 4).

**Figure 4.**
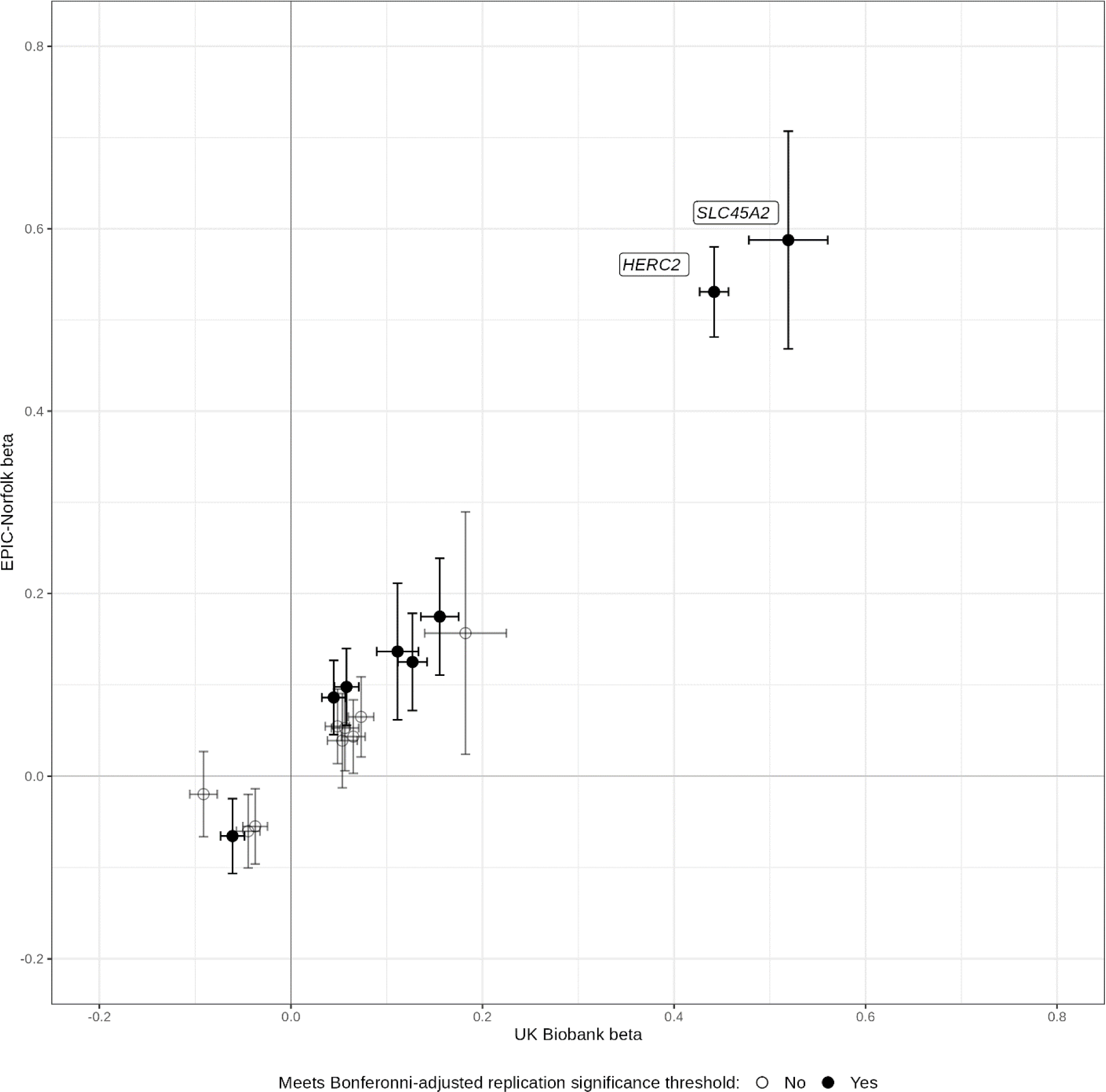
Comparison of betas expressed as change in standard deviation of mean RPS for lead variants identified from the discovery (UK Biobank) genome-wide association study (GWAS) with their corresponding betas in the replication (EPIC-Norfolk) analysis, with 95% confidence intervals. Variants meeting the Bonferroni-adjusted replication significance threshold (p = 0·05/17 variants) in the EPIC-Norfolk GWAS are shaded black. The nearest gene is annotated for variants achieving genome-wide significance.

### Phenome-Wide Association Study

A PheWAS was performed within the discovery UK Biobank GWAS sub-cohort (n=37,067) to assess potential associations between RPS with 308 diseases. After correction for multiple testing (p<0·05/308), significant associations were observed for higher RPS (indicating more pigmentation of the retina) with decreased odds of ‘Actinic keratosis’ (OR: 0·87, 95%CI: [0·81, 0·93]), ‘Primary Malignancy-Other Skin and subcutaneous tissue’ (0·90 [0·86, 0·94]), and ‘Migraine’ (0·91, [0·87, 0·95]). Higher RPS was associated with greater risk of chronic obstructive pulmonary disease; (1·11, [1·05, 1·16]). A further 26 diseases were significantly associated at p<0·05, including decreased odds for ‘Primary Malignancy-Malignant Melanoma’ (p 0·02) (supplementary figure 4, supplementary table 5).

#### Mendelian randomization

Two-sample MR analyses were performed to probe potential causal relationships between genetically predicted retinal pigmentation with outcomes of particular interest, as highlighted by the PheWAS analysis, using outcome summary statistics from Finngen[29] (supplementary table 6). MR estimates (OR per SD change in RPS [95%CI]) provided evidence for protective causal effects on actinic keratosis (0·44, [0·24, 0·83]; p 0·01), basal cell carcinoma of the skin (0·59, [0·38, 0·92]; p 0·02), squamous cell carcinoma of the skin (0·38, [0·20, 0·73]; p 0·003), non-melanoma skin cancer (0·40, [0·22, 0·73]; p 0·03) and malignant melanoma of the skin (0·60, [0·38, 0·94]; p 0·003).

## Discussion

We introduce a metric, the RPS, which quantifies the background pigmentation of the retina from colour photographs along a continuous scale and is strongly associated with genetic variants linked to human skin, eye, and hair phenotypes with replication in an additional cohort. We also found that clinical variables such as height and refractive error were associated with RPS. The RPS captures the biological variability of retinal colour without recourse to the distinct, social and political constructs of race and ethnicity. There is a significant overlap in the distribution of RPS among all ethnic groups and a wide range of RPS for each ethnicity (figure 2). Other than Chinese participants (a relatively small cohort in this dataset), each ethnicity has RPS scores that fall within each quintile of the RPS distribution.

The Fitzpatrick classification of skin types (FST) scale has been adopted in computer science to describe diversity in imaging datasets and exposes underlying biases within AI algorithms. Studies have found that facial recognition AI performs worse on individuals with higher FST (darker skin colour)[12] and object detection software is worse at detecting pedestrians with higher skin phototypes from street traffic-derived images.[11] This led the Google Ethical AI team to recommend that all computer vision models report their performance across the range of FST.[30]

There is some evidence that retinal colour affects model performance. A deep learning model trained to predict AMD found that patients with the minor allele at the rs12913832/*HERC2* locus, which was found in our study to be associated with retinal pigmentation, were more likely to have false positives for AMD.[31] Additionally, saturation values from retinal oximetry vary according to retinal pigmentation.[32]

We found associations between the RPS and multiple genetic loci previously associated with skin, hair, and iris colour, suggesting that the RPS reflects the degree of retinal pigmentation. Of the 20 genome-wide significant loci identified by conditional analysis in the discovery GWAS analysis, 17 had previously studied associations with hair, skin or iris pigmentation, including 3 that are known to be associated with oculocutaneous albinism (*TYR*, *OCA2* and *TYRP1*).[33]. Furthermore despite differences in populations and fundus cameras used, we observed replication for these loci in the EPIC-Norfolk cohort and strong correlation between beta coefficients in the two cohorts. This suggests that despite a range of input data characteristics. the RPS is still estimating retinal pigmentation Post-GWAS analyses for a set of 100 prioritised causal genes demonstrated enrichment for various melanin and pigmentation pathways, as well as enrichment for pigmentation-related traits in the GWAS catalog.[28]

The two most significantly associated loci in the discovery GWAS analysis were at *HERC2* (rs12913832), and *SLC45A2* (rs16891982). These also reached genome-wide significance in the replication analysis. The former is known to influence melanin production via effects on OCA2 expression, and iris colour,[34,35] whilst rs16891982 is a missense mutation in the *SLC45A2* gene.[36] rs12913832 modulates human pigmentation by affecting chromatin-loop formation between a long-range enhancer and the *OCA2* promoter, leading to decreased expression of OCA2 and lighter pigmentation.[34] This variant is strongly associated with brown iris colour in European populations.[35] The *SLC45A2* gene encodes a membrane protein involved in the transport of solutes including tyrosine (a precursor to melanin synthesis), which is implicated in the regulation of skin, hair and iris colour.[37,38] rs16891982 encodes a missense mutation in *SCL45A2*, and has been associated with skin pigmentation as well as a strong association with risk for cutaneous malignant melanoma.[39]

Interestingly, the lead variants at *PDE3A*, *SIK1* and *IFT122* have not been previously associated with skin, hair or iris pigmentation, thus these new variants may be specifically related to retinal pigmentation. *PDE3A* has been previously associated with arteriolar tortuosity[40], *SIK1* with regulation of circadian rhythms[41], and in vitro work has implicated alternative splicing for *IFT122* to play a role in PRPF31 retinitis pigmentosa pathogenesis.[42]

Diseases associated with skin pigmentation were also associated with retinal pigmentation. Actinic keratosis and cutaneous malignancy were inversely associated with increased RPS at phenome-wide significance. Malignant melanoma was also inversely associated with increased RPS, albeit only nominally, possibly due to the limited number of participants (n=643) with malignant melanoma. MR analyses furthermore provided evidence that genetic predisposition to increased retinal pigmentation is causally protective for skin malignancies, including malignant melanoma.

The RPS may be utilised for several indications in the future. First, reporting RPS for AI training or test datasets could allow immediate description of the phenotypic diversity. Second, it could be used as a standard metric in evaluating differences in AI algorithm performance across a diverse set of input images similar to how FST has been used in dermatology. This has implications not just for ophthalmic diseases, but also other diseases using retinal imaging biomarkers, such as Alzheimer’s disease[43] and cardiovascular disease.[44] Limitations of our study are as follows. Both the UK Biobank and EPIC-Norfolk participants are predominantly self-reported white and European. However, 7·5% of UK Biobank participants equates to over 3,000 people reporting non-white ethnicity. The RPS method is open source to allow application to other datasets to address this limitation. Secondly, RPS is currently dataset specific, so that absolute RPS values from different cohorts cannot be directly compared. This may be resolved with standardisation of the metric between camera types, using device specific raw RGB values. Finally, the performance of the RPS in retinal disease states will need to be assessed in future work.

In conclusion, the RPS is a continuous metric of retinal pigmentation derived directly from a retinal image with associations with genes implicated in hair, eye, and skin colour. The RPS is more task specific than ethnicity to estimate retinal pigmentation. This may have implications for AI algorithm development, testing, and for inclusion and algorithmic fairness across all fields of medicine that use retinal imaging as a biomarker.

## Data Availability

The code is available on github: https://github.com/uw-biomedical-ml/retinal-pigmentation-score

## Author contributions

CE, AYL, AER, YW, and AO-B designed the study. AER, YW, AO-B, ANW, KS, MB, CYU, RL undertook image analysis, traditional statistics, and genetic analyses. AER, AYL, AO-B, and ANW had direct access, and verified, the underlying data. AER, AO-B, AYL, ANW, and CE wrote the first version of the manuscript. All authors, interpreted results, critically reviewed, and edited the manuscript. AO-B had the final responsibility for the decision to submit for publication. All authors had access to the data presented and have approved the decision to submit for publication.

## Data availability statement

UK Biobank data are made available to approved researchers through a procedure described at http://www.ukbiobank.ac.uk/using-the-resource/. Retinal pigment scores for UK Biobank participants will be made available to approved UK Biobank researchers as a returned dataset (https://biobank.ndph.ox.ac.uk/ukb/docs.cgi?id=1). Finngen genome-wide association study (GWAS) summary statistics are publicly available online (https://www.finngen.fi/en/access_results). Summary statistics from the GWAS analyses presented in this study will be made publicly available from the NHGRI-EBI Catalog of human genome-wide association studies (https://www.ebi.ac.uk/gwas/).

## Code availability statement

The code to derive RPS is publicly available at: https://github.com/uw-biomedical-ml/retinal-pigmentation-score.

## Acknowledgements

The authors would like to thank the 500,000 UK Biobank participants, the participants and the investigators of the FinnGen study and the EPIC-Norfolk study, and the funders of these projects.

C.E., and A.T. received a proportion of their financial support from the UK Department of Health through an award made by the National Institute for Health Research to Moorfields Eye Hospital NHS Foundation Trust and UCL Institute of Ophthalmology for a Biomedical Research Centre for Ophthalmology.

C.S.L receives grant funding from NIH/NIA R01AG060942, U19AG066567, OT2OD032644, and the Klorfine Family Endowed Chair, and the Karalis Johnson Retina Center.

A.Y.L. has received an unrestricted and career development award from Research to Prevent Blindness, Latham Vision Science Awards, NEI/NIH K23EY029246, OT2OD032644, the C. Dan and Irene Hunter Endowed Professorship, and the Karalis Johnson Retina Center.

APK is supported by a UK Research and Innovation Future Leaders Fellowship, an Alcon Research Institute Young Investigator Award and a Lister Institute for Preventive Medicine Award. This research was supported by the NIHR Biomedical Research Centre at Moorfields Eye Hospital and the UCL Institute of Ophthalmology.

