## Supplemental Methods and Figures for "Ethnicity is not biology: retinal pigment score to evaluate biological variability from ophthalmic imaging using machine learning"

### Table of contents

Supplementary material 3

Supplementary methods 3

GWAS analysis 3

Mendelian randomisation 4

Supplementary results 5

Supplementary Table 1. Baseline cohort characteristics by tertiles of retinal pigment score (RPS). 5

Supplementary Table 2: Multivariable Linear regression results. 6

Supplementary Table 3: Prioritised gene set from the discovery UK Biobank genome-wide association study. 7

Supplementary Table 4: EPIC-Norfolk GWAS results 11

Supplementary Table 5: PheWAS table of results 13

Supplementary Table 6. Details of summary-level data used for Mendelian randomization analyses. 16

Supplemental Table 7. Mendelian randomization results for potentially causal associations between retinal pigment score with selected outcomes of interest. 17

Supplementary Table 8. Tests of heterogeneity, directional pleiotropy and regression dilution statistics for the retinal pigment score instrumental variable 18

Supplementary Figure 1: Adjusted Regression plots. Adjusted mean retinal pigment score by covariates of interest. Adjusted means (solid black dots), 95% confidence intervals (vertical solid lines), and regression line for continuous variables (dotted line) are from a linear regression model adjusting for age, sex, and UK biobank centre. *M, myopia; E, emmetropia; H, hyperopia. 19

Supplementary Figure 2. Traits that are significantly enriched for prioritised genes from the discovery genome-wide association study, as evidenced by previously reported gene-trait associations in the GWAS Catalog. 20

Supplementary Figure 3. Gene Ontology biological pathways from MsigDB significantly enriched for prioritised genes from the discovery genome-wide association study. 20

Supplementary Figure 4: PheWAS Volcano Plot 21

References 22

### Supplementary material

#### Supplementary methods

Details of the overall UK Biobank study protocol and protocols for individual tests are available online (https://biobank.ndph.ox.ac.uk/ukb/index.cgi).

The choices for ethnic background in UK Biobank were categorised as white, Mixed, Asian or Asian British, black or black British, Chinese, or other ethnic group. (https://biobank.ndph.ox.ac.uk/ukb/index.cgi).

The European Prospective Investigation into Cancer and Nutrition-Norfolk (EPIC-Norfolk) Eye Study protocol is available online at https://www.epic-norfolk.org.uk/.

##### GWAS analysis

Full details for genotyping and imputation in the UK Biobank cohort have been described previously.[1] In brief, genotype calling was performed using two arrays: the UK BiLEVE Axiom array (~50,000 participants) and the UK Biobank Axiom array (~450,000 participants). Marker positions are in GRCh37 coordinates. There were 805,426 markers available in the released data after quality control. Genotype imputation was then performed using a combined Haplotype Reference Consortium and UK10K reference panel, expanding the number of testable variants to ~96 million. The majority of the EPIC-Norfolk cohort were also genotyped using the Affymetrix UK Biobank Axiom Array, however, genotyping for a small subset was undertaken using the Affymetrix GeneChip Human Mapping 500K Array Set.

The following exclusions were applied for sample quality control: individuals with relatedness corresponding to third-degree relatives or closer, excess of missing genotypes or more heterozygosity than expected. The GWAS analysis was furthermore restricted to individuals of European ethnicity only. The following exclusions were applied for variant-level quality control: call rate <95%, Hardy-Weinberg equilibrium $p<1x10^{-6}$, posterior call probability <0.9, INFO score <0.9 and minor allele frequency <0.01.

The SNP2GENE function within FUMA[2] was used to perform positional and expression quantitative trait locus (eQTL) gene mapping/prioritisation. SNPs in high LD ($r^2>0.6$) with any independent lead variant were positionally mapped to genes located within 10kb. Variants were also mapped to a set of prioritised genes within 1Mb if associated with the expression of those genes in retina (reported in EyeGEx), skin (reported in TwinsUK, GTEx v8) and dermal fibroblasts (reported in GTEx v8). To test the prioritised genes for enrichment in biological pathways, FUMA's GENE2FUNC function was applied, using hypergeometric mean pathway analysis against gene sets obtained from MsigDB and WikiPathways. Multiple testing correction (Benjamini-Hochberg method) per data source of tested gene sets was performed and gene sets with adjusted P≤0.05 and >1 overlapping genes were reported by default.

The GWAS analysis was performed using REGENIE software.[3] GCTA-COJO was used to prioritise lead variants.[4] Genomic inflation factor and heritability estimates were calculated using the LDSC tool[5] and pre-calculated LD scores for European ancestry (https://data.broadinstitute.org/alkesgroup/LDSCORE/eur_w_ld_chr.tar.bz2). All other statistical analyses and were performed in R (R for GNU macOS, Version 4.2.0, The R Foundation for Statistical Computing, Vienna, Austria).[6] R packages used included PheWAS[7], TwoSampleMR[8], MendelianRandomization[9], ukbwranglr[10], codemapper[11], targets[12], tarchetypes[13], tidyverse [14], workflowr[15], flextable[16], gtsummary[17] and knitr.[18]

#### Mendelian randomisation

Two-sample Mendelian randomisation (MR) analyses were conducted drawing on the discovery RPS GWAS results (exposure) and summary statistics from the independent Finngen cohort[19] (data release 8) for selected outcomes of interest, as highlighted by the RPS PheWAS analysis. The IV for retinal pigmentation was constructed by selecting all conditionally independent genome-wide significant single nucleotide polymorphisms (SNPs) from the discovery RPS GWAS analysis, and performing clumping to exclude those with linkage disequilibrium $R^2>0.01$ and within 10 000 kb, using the 1000 Genomes Project European reference population.[20] Palindromic SNPs with minor allele frequency >0.46 were excluded. Effect alleles were harmonized across exposure and outcome datasets.

The multiplicative random-effects inverse-variance weighted (IVW) approach method used for MR analyses provides precise and efficient estimates, but is sensitive to invalid instrumental variables (IVs) and pleiotropy.[30] We therefore conducted sensitivity analyses using three alternative MR methods: weighted median[31], weighted mode[32] and MR-Egger[33]. Each method makes different assumptions about the nature of pleiotropy and consistent estimates across methods strengthens causal inferences.[34]

Under the IVW method, we calculated the mean F-statistic as an indicator of instrument strength (a value >10 is usually considered a strong instrument).[21] We assessed for heterogeneity with the $I^2$ and Cochran's $Q$ statistics in the IVW model and with Rucker's $Q'$ statistic in MR-Egger regression. The $I^2_{GX}$ statistic is an indicator of expected relative bias (or dilution) of the MR-Egger causal estimate.[22] In MR-Egger regression, a significant difference of the intercept from zero is evidence for average directional horizontal pleiotropy.[23]

### Supplementary results

#### Supplementary Table 1. Baseline cohort characteristics by tertiles of retinal pigment score (RPS).

| | | RPS tertiles | | |
|---|---|---|---|---|
| **Characteristic** | **Overall**, N = 44,320 | **1**, N = 14,774 | **2**, N = 14,773 | **3**, N = 14,773 |
| **Retinal pigment score** | -0.82 (-9.89, 10.39) | -12.64 (-15.68, -9.89) | -0.82 (-4.12, 2.59) | 15.18 (10.39, 21.43) |
| **Age** | 56 (49, 63) | 56 (49, 62) | 58 (50, 63) | 55 (48, 62) |
| **Sex** | | | | |
| Female | 24,414 (55%) | 8,109 (55%) | 8,182 (55%) | 8,123 (55%) |
| Male | 19,906 (45%) | 6,665 (45%) | 6,591 (45%) | 6,650 (45%) |
| **Ethnicity*** | | | | |
| White | 40,704 (92%) | 14,679 (100%) | 14,551 (99%) | 11,474 (78%) |
| Black | 1,135 (2.6%) | 1 (<0.1%) | 9 (<0.1%) | 1,125 (7.7%) |
| Asian | 1,078 (2.4%) | 5 (<0.1%) | 37 (0.3%) | 1,036 (7.1%) |
| Mixed | 369 (0.8%) | 11 (<0.1%) | 42 (0.3%) | 316 (2.2%) |
| Chinese | 161 (0.4%) | 0 (0%) | 2 (<0.1%) | 159 (1.1%) |
| Other | 599 (1.4%) | 20 (0.1%) | 64 (0.4%) | 515 (3.5%) |
| **Skin color*** | | | | |
| Very fair | 3,583 (8.2%) | 1,682 (11%) | 1,226 (8.4%) | 675 (4.7%) |
| Fair | 28,889 (66%) | 11,044 (75%) | 10,356 (71%) | 7,489 (52%) |
| Light olive | 8,219 (19%) | 1,772 (12%) | 2,759 (19%) | 3,688 (26%) |
| Dark olive | 779 (1.8%) | 114 (0.8%) | 202 (1.4%) | 463 (3.2%) |
| Brown | 1,705 (3.9%) | 23 (0.2%) | 69 (0.5%) | 1,613 (11%) |
| Black | 482 (1.1%) | 0 (0%) | 1 (<0.1%) | 481 (3.3%) |
| **Hair color*** | | | | |
| Blonde | 4,567 (10%) | 2,317 (16%) | 1,513 (10%) | 737 (5.0%) |
| Red | 1,781 (4.0%) | 701 (4.8%) | 644 (4.4%) | 436 (3.0%) |
| Light brown | 16,657 (38%) | 6,564 (45%) | 6,103 (41%) | 3,990 (27%) |
| Dark brown | 16,656 (38%) | 4,572 (31%) | 5,641 (38%) | 6,443 (44%) |
| Black | 3,980 (9.0%) | 393 (2.7%) | 648 (4.4%) | 2,939 (20%) |
| Other | 486 (1.1%) | 182 (1.2%) | 174 (1.2%) | 130 (0.9%) |
| **Height (in cm)** | 168 (162, 176) | 169 (163, 176) | 168 (162, 176) | 168 (162, 175) |
| **Townsend index of deprivation (quintiles)*** | | | | |
| 1 (least deprived) | 13,047 (29%) | 4,723 (32%) | 4,564 (31%) | 3,760 (25%) |
| 2 | 9,029 (20%) | 3,138 (21%) | 3,062 (21%) | 2,829 (19%) |
| 3 | 8,447 (19%) | 2,796 (19%) | 2,844 (19%) | 2,807 (19%) |
| 4 | 8,303 (19%) | 2,665 (18%) | 2,693 (18%) | 2,945 (20%) |
| 5 (more deprived) | 5,429 (12%) | 1,427 (9.7%) | 1,591 (11%) | 2,411 (16%) |

Median (IQR) for continuous variables.
Count (column %) for categorical variables.
*Variables with missing data: ethnicity (n=274, 0.6%), skin color (n=663, 1.5%), hair color (n=193, 0.4%), Townsend index of deprivation (n=65, 0.1%)

Supplementary Table 2: Multivariable Linear regression results.

| Characteristic | Beta | 95% CI | p-value |
|---|---|---|---|
| **Age (per 5 years)** | 0.20 | 0.13, 0.27 | **1.3e-08** |
| **Sex** | | | |
| Female | — | — | |
| Male | 0.17 | -0.14, 0.48 | 0.279 |
| **Ethnicity** | | | |
| White | — | — | |
| Black | 15 | 14, 16 | **1.6e-158** |
| Asian | 14 | 14, 15 | **4.0e-216** |
| Mixed | 12 | 11, 13 | **4.7e-86** |
| Chinese | 20 | 18, 21 | **4.6e-103** |
| Other | 13 | 12, 14 | **2.8e-135** |
| Missing | 5.8 | 4.3, 7.4 | **1.4e-13** |
| **Skin color** | | | |
| Very fair | — | — | |
| Fair | 2.2 | 1.8, 2.6 | **5.9e-27** |
| Light olive | 6.3 | 5.8, 6.7 | **8.2e-147** |
| Dark olive | 7.3 | 6.4, 8.2 | **4.3e-55** |
| Brown | 9.5 | 8.5, 10 | **4.3e-88** |
| Black | 11 | 9.4, 12 | **5.7e-48** |
| Missing | 6.2 | 5.1, 7.2 | **5.2e-32** |
| **Hair color** | | | |
| Blonde | — | — | |
| Red | 3.5 | 2.9, 4.1 | **4.1e-28** |
| Light brown | 2.1 | 1.7, 2.5 | **5.1e-29** |
| Dark brown | 4.9 | 4.5, 5.3 | **1.2e-136** |
| Black | 7.0 | 6.4, 7.6 | **4.8e-122** |
| Other | 2.3 | 1.2, 3.3 | **1.9e-05** |
| Missing | 3.6 | 1.7, 5.5 | **2.0e-04** |
| **Refractive status** | | | |
| Myopia <-6.00D | — | — | |
| Myopia | 1.6 | 1.1, 2.2 | **1.3e-08** |
| Emmetropia | 2.1 | 1.5, 2.6 | **1.1e-12** |
| Hyperopia | 1.4 | 0.84, 2.0 | **1.1e-06** |
| Hyperopia >+6.00D | -1.3 | -2.9, 0.38 | 0.133 |
| **Height (per 5cm)** | -0.24 | -0.32, -0.15 | **3.6e-08** |
| **Townsend index of deprivation (quintiles)** | | | |
| 1 (least deprived) | — | — | |
| 2 | 0.11 | -0.19, 0.41 | 0.480 |
| 3 | 0.15 | -0.16, 0.46 | 0.353 |
| 4 | 0.22 | -0.10, 0.54 | 0.169 |
| 5 (more deprived) | 0.81 | 0.43, 1.2 | **2.5e-05** |
| Missing | -0.30 | -3.0, 2.5 | 0.833 |

Differences in retinal pigment score (RPS) per specified differences in covariates. Estimates are mutually adjusted for all covariates shown in the table. Bold p-values mean statistically significant results.

Supplementary Table 3: Prioritised gene set from the discovery UK Biobank genome-wide association study.

| Ensembl ID | Gene symbol |
|---|---|
| ENSG00000198625 | *MDM4* |
| ENSG00000184144 | *CNTN2* |
| ENSG00000174529 | *TMEM81* |
| ENSG00000117222 | *RBBP5* |
| ENSG00000133059 | *DSTYK* |
| ENSG00000133069 | *TMCC2* |
| ENSG00000163545 | *NUAK2* |
| ENSG00000162873 | *KLHDC8A* |
| ENSG00000066027 | *PPP2R5A* |
| ENSG00000065600 | *TMEM206* |
| ENSG00000117691 | *NENF* |
| ENSG00000143801 | *PSEN2* |
| ENSG00000163050 | *ADCK3* |
| ENSG00000172771 | *EFCAB12* |
| ENSG00000129071 | *MBD4* |
| ENSG00000163913 | *IFT122* |
| ENSG00000163914 | *RHO* |
| ENSG00000178804 | *H1FOO* |
| ENSG00000004399 | *PLXND1* |
| ENSG00000151388 | *ADAMTS12* |
| ENSG00000182631 | *RXFP3* |
| ENSG00000164175 | *SLC45A2* |
| ENSG00000137265 | *IRF4* |
| ENSG00000112685 | *EXOC2* |
| ENSG00000188996 | *HUS1B* |
| ENSG00000028839 | *TBPL1* |

| | |
|---|---|
| ENSG00000146411 | *SLC2A12* |
| ENSG00000196367 | *TRRAP* |
| ENSG00000198742 | *SMURF1* |
| ENSG00000185467 | *KPNA7* |
| ENSG00000106245 | *BUD31* |
| ENSG00000106246 | *PTCD1* |
| ENSG00000248919 | *ATP5J2-PTCD1* |
| ENSG00000198556 | *ZNF789* |
| ENSG00000160908 | *ZNF394* |
| ENSG00000196652 | *ZKSCAN5* |
| ENSG00000021461 | *CYP3A43* |
| ENSG00000106261 | *ZKSCAN1* |
| ENSG00000166529 | *ZSCAN21* |
| ENSG00000166526 | *ZNF3* |
| ENSG00000168090 | *COPS6* |
| ENSG00000166508 | *MCM7* |
| ENSG00000221838 | *AP4M1* |
| ENSG00000106290 | *TAF6* |
| ENSG00000197093 | *GAL3ST4* |
| ENSG00000213420 | *GPC2* |
| ENSG00000066923 | *STAG3* |
| ENSG00000160844 | *GATS* |
| ENSG00000214300 | *SPDYE3* |
| ENSG00000121716 | *PILRB* |
| ENSG00000085514 | *PILRA* |
| ENSG00000078487 | *ZCWPW1* |
| ENSG00000146834 | *MEPCE* |
| ENSG00000160813 | *PPP1R35* |
| ENSG00000185955 | *C7orf61* |
| ENSG00000166925 | *TSC22D4* |

| | |
|---|---|
| ENSG00000166924 | *NYAP1* |
| ENSG00000077080 | *ACTL6B* |
| ENSG00000172354 | *GNB2* |
| ENSG00000146830 | *GIGYF1* |
| ENSG00000196411 | *EPHB4* |
| ENSG00000146828 | *SLC12A9* |
| ENSG00000087087 | *SRRT* |
| ENSG00000087085 | *ACHE* |
| ENSG00000205277 | *MUC12* |
| ENSG00000169876 | *MUC17* |
| ENSG00000128581 | *RABL5* |
| ENSG00000107165 | *TYRP1* |
| ENSG00000153714 | *LURAP1L* |
| ENSG00000110693 | *SOX6* |
| ENSG00000110075 | *PPP6R3* |
| ENSG00000132749 | *MTL5* |
| ENSG00000197345 | *MRPL21* |
| ENSG00000162341 | *TPCN2* |
| ENSG00000172927 | *MYEOV* |
| ENSG00000123892 | *RAB38* |
| ENSG00000168959 | *GRM5* |
| ENSG00000077498 | *TYR* |
| ENSG00000086991 | *NOX4* |
| ENSG00000172572 | *PDE3A* |
| ENSG00000134532 | *SOX5* |
| ENSG00000080166 | *DCT* |
| ENSG00000152749 | *GPR180* |
| ENSG00000104044 | *OCA2* |
| ENSG00000128731 | *HERC2* |
| ENSG00000183629 | *GOLGA8G* |

| | |
|---|---|
| ENSG00000188626 | *GOLGA8M* |
| ENSG00000034053 | *APBA2* |
| ENSG00000184009 | *ACTG1* |
| ENSG00000186765 | *FSCN2* |
| ENSG00000185504 | *C17orf70* |
| ENSG00000182446 | *NPLOC4* |
| ENSG00000185527 | *PDE6G* |
| ENSG00000204237 | *OXLD1* |
| ENSG00000185298 | *CCDC137* |
| ENSG00000214087 | *ARL16* |
| ENSG00000185359 | *HGS* |
| ENSG00000262814 | *MRPL12* |
| ENSG00000262660 | *SLC25A10* |
| ENSG00000183048 | *SLC25A10* |

Supplementary Table 4: EPIC-Norfolk GWAS results

| Rs identifier | chr:pos [hg19] | EA/OA (EAF) | Beta (95% CI) | P | Nearest gene | Genome-wide significant |
|---|---|---|---|---|---|---|
| rs6670870 | 1:205155177 | A/T (0.76) | -0.02 (-0.07; 0.03) | 4.1E-01 | *DSTYK* | |
| rs173273 | 1:212446689 | G/T (0.41) | – | – | *PPP2R5A* | |
| rs762948237 | 3:129178587 | TCTTC/T (0.87) | – | – | *IFT122* | |
| rs16891982 | 5:33951693 | C/G (0.02) | 0.59 (0.47; 0.71) | 5.1E-22 | *SLC45A2* | Yes |
| rs12203592 | 6:396321 | C/T (0.79) | 0.13 (0.07; 0.18) | 4.2E-06 | *IRF4* | |
| rs62425803 | 6:134330249 | G/A (0.81) | 0.04 (-0.01; 0.09) | 1.4E-01 | *TCF21* | |
| rs117756744 | 7:100277212 | G/A (0.98) | 0.16 (0.02; 0.29) | 2.1E-02 | *GNB2* | |
| rs1325117 | 9:12613472 | G/A (0.36) | 0.1 (0.05; 0.14) | 5.0E-06 | *TYRP1;LURAP1L* | |
| rs11023814 | 11:16007053 | C/G (0.43) | 0.09 (0.04; 0.13) | 3.1E-05 | *SOX6* | |
| rs150527451 | 11:68817897 | G/A (0.89) | 0.17 (0.11; 0.24) | 8.1E-08 | *TPCN2* | |
| rs1060435 | 11:68855595 | A/G (0.59) | 0.04 (0; 0.08) | 3.5E-02 | *TPCN2* | |
| rs747572 | 11:87885082 | A/G (0.63) | 0.05 (0.01; 0.1) | 8.6E-03 | *CTSC* | |
| rs1126809 | 11:89017961 | G/A (0.7) | 0.06 (0.02; 0.11) | 3.7E-03 | *TYR* | |

| | | | | | | | |
|---|---|---|---|---|---|---|---|
| rs4762973 | 12:20710145 | A/G (0.75) | 0.05 (0; 0.1) | 2.8E-02 | *PDE3A* | | |
| rs10771034 | 12:23979199 | T/A (0.45) | -0.06 (-0.1; -0.02) | 3.3E-03 | *SOX5* | | |
| rs766338951 | 13:95169060 | CT/C (0.69) | – | – | *DCT* | | |
| rs1800407 | 15:28230318 | C/T (0.91) | 0.14 (0.06; 0.21) | 3.4E-04 | *OCA2* | | |
| rs12913832 | 15:28365618 | A/G (0.22) | 0.53 (0.48; 0.58) | 1.2E-98 | *HERC2* | | Yes |
| rs7220155 | 17:79606020 | C/T (0.62) | -0.07 (-0.11; -0.02) | 1.7E-03 | *TSPAN10* | | |
| rs1785433 | 21:44783282 | A/G (0.65) | -0.05 (-0.1; -0.01) | 9.0E-03 | *SIK1* | | |

#### Supplementary Table 5: PheWAS table of results

*Table of retinal pigment score phenome-wide association study results meeting the nominal significance threshold (p<0.05).*

| category | phenotype | n_cases | n_controls | p | OR (95% CI) |
|---|---|---|---|---|---|
| **Benign Neoplasm/CIN** | | | | | |
| | Benign neoplasm and polyp of uterus | 1,042 | 36,025 | 0.0230 | 1.08 (1.01; 1.15) |
| **Cancers** | | | | | |
| | Primary Malignancy_Other Skin and subcutaneous tissue | 2,334 | 34,733 | 0.0000 | 0.9 (0.86; 0.94) |
| | Primary Malignancy_Malignant Melanoma | 644 | 36,423 | 0.0172 | 0.91 (0.83; 0.98) |
| **Circulatory System** | | | | | |
| | Myocardial infarction | 1,482 | 35,585 | 0.0165 | 1.07 (1.01; 1.13) |
| | Venous thromboembolic disease (Excl PE) | 1,095 | 35,972 | 0.0232 | 1.07 (1.01; 1.14) |
| **Digestive System** | | | | | |
| | Gastro-oesophageal reflux disease | 5,622 | 31,445 | 0.0015 | 0.95 (0.93; 0.98) |
| | Abdominal Hernia | 4,355 | 32,712 | 0.0027 | 1.05 (1.02; 1.09) |
| | Diverticular disease of intestine (acute and chronic) | 4,484 | 32,583 | 0.0198 | 1.04 (1.01; 1.07) |
| **Eye** | | | | | |
| | Diabetic ophthalmic complications | 629 | 36,438 | 0.0293 | 0.91 (0.84; 0.99) |
| | Anterior and Intermediate Uveitis | 205 | 36,862 | 0.0324 | 0.86 (0.74; 0.99) |

| | | | | | |
|---|---|---|---|---|---|
| | Retinal detachments and breaks | 449 | 36,618 | 0.0480 | 0.91 (0.82; 1) |
| **Genitourinary system** | | | | | |
| | Postcoital and contact bleeding | 323 | 36,744 | 0.0315 | 0.88 (0.79; 0.99) |
| | Erectile dysfunction | 1,201 | 35,866 | 0.0363 | 0.94 (0.88; 1) |
| **Haematological/Immunological conditions** | | | | | |
| | Agranulocytosis | 490 | 36,577 | 0.0020 | 0.86 (0.78; 0.95) |
| **Infectious Diseases** | | | | | |
| | Infection of skin and subcutaneous tissues | 1,462 | 35,605 | 0.0004 | 1.1 (1.04; 1.16) |
| | Lower Respiratory Tract Infections | 3,035 | 34,032 | 0.0024 | 1.06 (1.02; 1.1) |
| | Bacterial Diseases (excl TB) | 5,091 | 31,976 | 0.0090 | 1.04 (1.01; 1.07) |
| | Other or unspecified infectious organisms | 4,922 | 32,145 | 0.0219 | 1.04 (1; 1.07) |
| **Mental Health Disorders** | | | | | |
| | Schizophrenia, schizotypal and delusional disorders | 154 | 36,913 | 0.0026 | 1.26 (1.08; 1.47) |
| **Musculoskeletal conditions** | | | | | |
| | Fracture of hip | 314 | 36,753 | 0.0062 | 1.17 (1.04; 1.31) |
| | Spondylolisthesis | 213 | 36,854 | 0.0063 | 1.2 (1.05; 1.38) |
| | Carpal tunnel syndrome | 1,673 | 35,394 | 0.0252 | 1.06 (1.01; 1.11) |

| | | | | | |
|---|---|---|---|---|---|
| **Neurological conditions** | | | | | |
| | Migraine | 2,462 | 34,605 | 0.0000 | 0.91 (0.87; 0.95) |
| **Respiratory System** | | | | | |
| | COPD | 1,664 | 35,403 | 0.0001 | 1.11 (1.05; 1.16) |
| | Allergic and chronic rhinitis | 4,310 | 32,757 | 0.0002 | 0.94 (0.91; 0.97) |
| | Chronic sinusitis | 2,992 | 34,075 | 0.0073 | 0.95 (0.91; 0.99) |
| | Pulmonary collapse (excl pneumothorax) | 450 | 36,617 | 0.0417 | 1.1 (1; 1.21) |
| **Skin conditions** | | | | | |
| | Actinic keratosis | 1,058 | 36,009 | 0.0000 | 0.87 (0.81; 0.93) |
| | Rosacea | 857 | 36,210 | 0.0019 | 0.9 (0.83; 0.96) |
| | Seborrheic dermatitis | 967 | 36,100 | 0.0327 | 0.93 (0.87; 1) |

Supplementary Table 6. Details of summary-level data used for Mendelian randomization analyses.

| Trait | Source | Description | Participants (cases/controls) |
|---|---|---|---|
| **Exposure** | | | |
| Retinal pigment score | UK Biobank | - | 37,067 |
| **Outcome** | | | |
| ***Dermatological*** | | | |
| Actinic keratosis | FinnGen | finngen_R8_L12_ACTINKERA | (9 319 / 331 962) |
| Basal cell carcinoma of skin | FinnGen | finngen_R8_C3_BASAL_CELL_CARCINOMA_EXALLC | (16 328 / 259 583) |
| Squamous cell carcinoma of skin | FinnGen | finngen_R8_C3_SQUAMOUS_CELL_CARCINOMA_SKIN_EXALLC | (2 749 / 259 583) |
| Malignant melanoma of skin | FinnGen | finngen_R8_C3_MELANOMA_SKIN_EXALLC | (2 705 / 259 583) |
| Other non-melanoma skin cancer | FinnGen | finngen_R8_C3_OTHER_SKIN_EXALLC | (14 863 / 259 583) |
| ***Other*** | | | |
| COPD | FinnGen | finngen_R8_J10_COPD | (16 410 / 283 589) |
| Migraine | FinnGen | finngen_R8_G6_MIGRAINE | (15 905 / 264 662) |

Abbreviations: SNP = single nucleotide polymorphism; IV = instrumental variable; COPD = chronic obstructive pulmonary disease.

Supplemental Table 7. Mendelian randomization results for potentially causal associations between retinal pigment score with selected outcomes of interest.

| MR method | Actinic keratosis | | | | Basal cell carcinoma of skin | | | | Squamous cell carcinoma of skin | | | | Malignant melanoma of skin | | | | Non-melanoma skin cancer | | | | COPD | | | | Migraine | | | |
|---|---|---|---|---|---|---|---|---|---|---|---|---|---|---|---|---|---|---|---|---|---|---|---|---|---|---|---|---|
| | Odds ratio | 95% LL | 95% UL | P-value | Odds ratio | 95% LL | 95% UL | P-value | Odds ratio | 95% LL | 95% UL | P-value | Odds ratio | 95% LL | 95% UL | P-value | Odds ratio | 95% LL | 95% UL | P-value | Odds ratio | 95% LL | 95% UL | P-value | Odds ratio | 95% LL | 95% UL | P-value |
| IVW | 0.441 | 0.236 | 0.827 | **0.011** | 0.591 | 0.382 | 0.916 | **0.019** | 0.384 | 0.202 | 0.729 | **0.003** | 0.404 | 0.223 | 0.730 | **0.003** | 0.597 | 0.381 | 0.936 | **0.025** | 1.080 | 0.950 | 1.228 | 0.239 | 1.031 | 0.868 | 1.226 | 0.726 |
| Weighted median | 0.611 | 0.471 | 0.794 | **<0.001** | 0.650 | 0.544 | 0.776 | **<0.001** | 0.541 | 0.365 | 0.802 | **0.002** | 0.441 | 0.313 | 0.621 | **<0.001** | 0.664 | 0.549 | 0.804 | **<0.001** | 1.106 | 0.966 | 1.267 | 0.144 | 0.971 | 0.826 | 1.141 | 0.721 |
| Weighted mode | 0.631 | 0.501 | 0.794 | **0.002** | 0.689 | 0.571 | 0.831 | **0.002** | 0.483 | 0.320 | 0.728 | **0.005** | 0.441 | 0.314 | 0.620 | **<0.001** | 0.713 | 0.592 | 0.859 | **0.004** | 1.125 | 0.981 | 1.292 | 0.118 | 0.949 | 0.785 | 1.147 | 0.597 |
| MR-Egger | 0.282 | 0.092 | 0.863 | **0.027** | 0.434 | 0.201 | 0.935 | **0.033** | 0.264 | 0.083 | 0.848 | **0.025** | 0.447 | 0.150 | 1.330 | 0.148 | 0.448 | 0.202 | 0.992 | **0.048** | 1.197 | 0.969 | 1.481 | 0.096 | 1.006 | 0.742 | 1.363 | 0.971 |

* Odds ratios expressed per standard deviation increase in retinal pigment score.

Supplementary Table 8. Tests of heterogeneity, directional pleiotropy and regression dilution statistics for the retinal pigment score instrumental variable

| MR method | Actinic keratosis | | Basal cell carcinoma of skin | | Squamous cell carcinoma of skin | | Malignant melanoma of skin | | Other non-melanoma skin cancer | | COPD | | Migraine | |
|---|---|---|---|---|---|---|---|---|---|---|---|---|---|---|
| | Estimate | P-value | Estimate | P-value | Estimate | P-value | Estimate | P-value | Estimate | P-value | Estimate | P-value | Estimate | P-value |
| **IVW** | | | | | | | | | | | | | | |
| Cochran's $Q$ statistic | 280.3 (12) | <0.001 | 217.5 (12) | <0.001 | 87.4 (12) | <0.001 | 77.7 (12) | <0.001 | 211.7 (12) | <0.001 | 20.3 (12) | 0.062 | 37.7 (12) | <0.001 |
| $I^2$ statistic | 95.7 % | - | 94.5 % | - | 86.3% | - | 84.6 % | - | 94.3% | - | 40.8 % | - | 68.1 % | - |
| **MR-Egger** | | | | | | | | | | | | | | |
| Rucker's $Q'$ statistic | 259.1 (11) | <0.001 | 200.7 (11) | <0.001 | 83.1 (11) | <0.001 | 77.3 (11) | <0.001 | 198.3 (11) | <0.001 | 18.0 (11) | 0.082 | 37.5 (11) | <0.001 |
| $I^2_{GX}$ statistic | 94.9 % | - | 95.3 % | - | 94.8% | - | 95.5 % | - | 95.3% | - | 96.1 % | - | 96.2 % | - |
| Intercept | 0.046 | 0.343 | 0.032 | 0.336 | 0.038 | 0.449 | -0.011 | 0.823 | 0.030 | 0.389 | -0.011 | 0.236 | 0.003 | 0.839 |

Supplementary Figure 1: Adjusted Regression plots. Adjusted mean retinal pigment score by covariates of interest. Adjusted means (solid black dots), 95% confidence intervals (vertical solid lines), and regression line for continuous variables (dotted line) are from a linear regression model adjusting for age, sex, and UK biobank centre. *M, myopia; E, emmetropia; H, hyperopia.

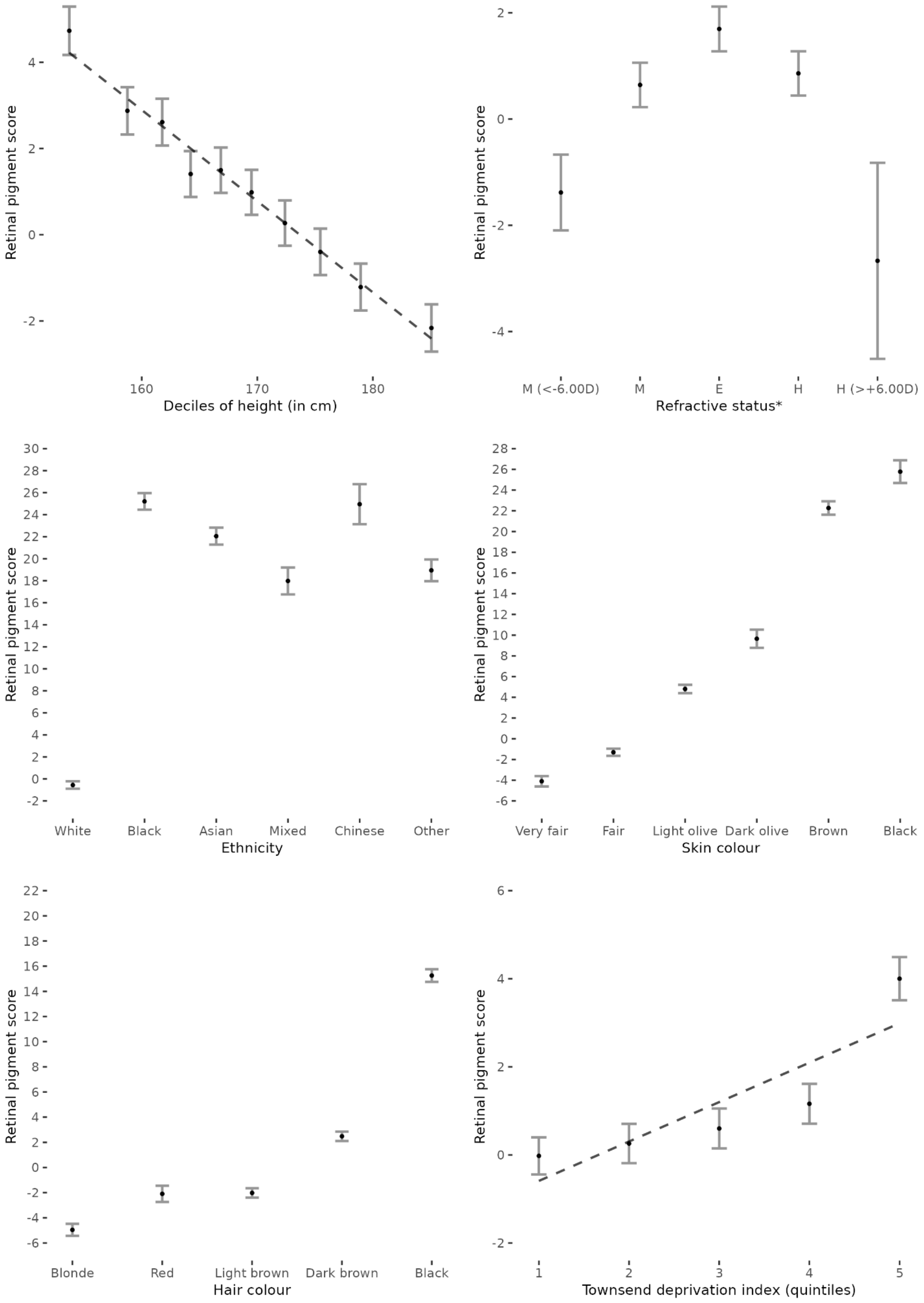

Supplementary Figure 2. Traits that are significantly enriched for prioritised genes from the discovery genome-wide association study, as evidenced by previously reported gene-trait associations in the GWAS Catalog.

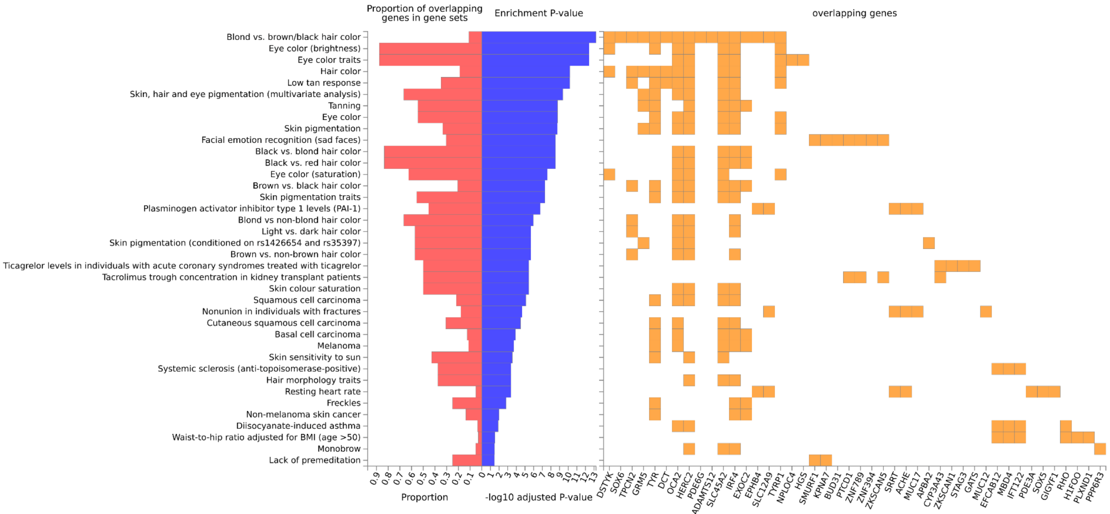


Supplementary Figure 3. Gene Ontology biological pathways from MsigDB significantly enriched for prioritised genes from the discovery genome-wide association study.

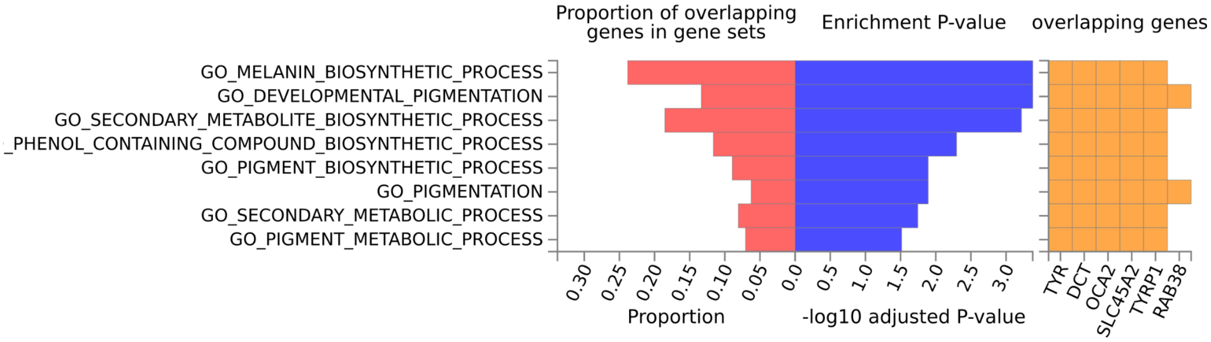

### Supplementary Figure 4: PheWAS Volcano Plot

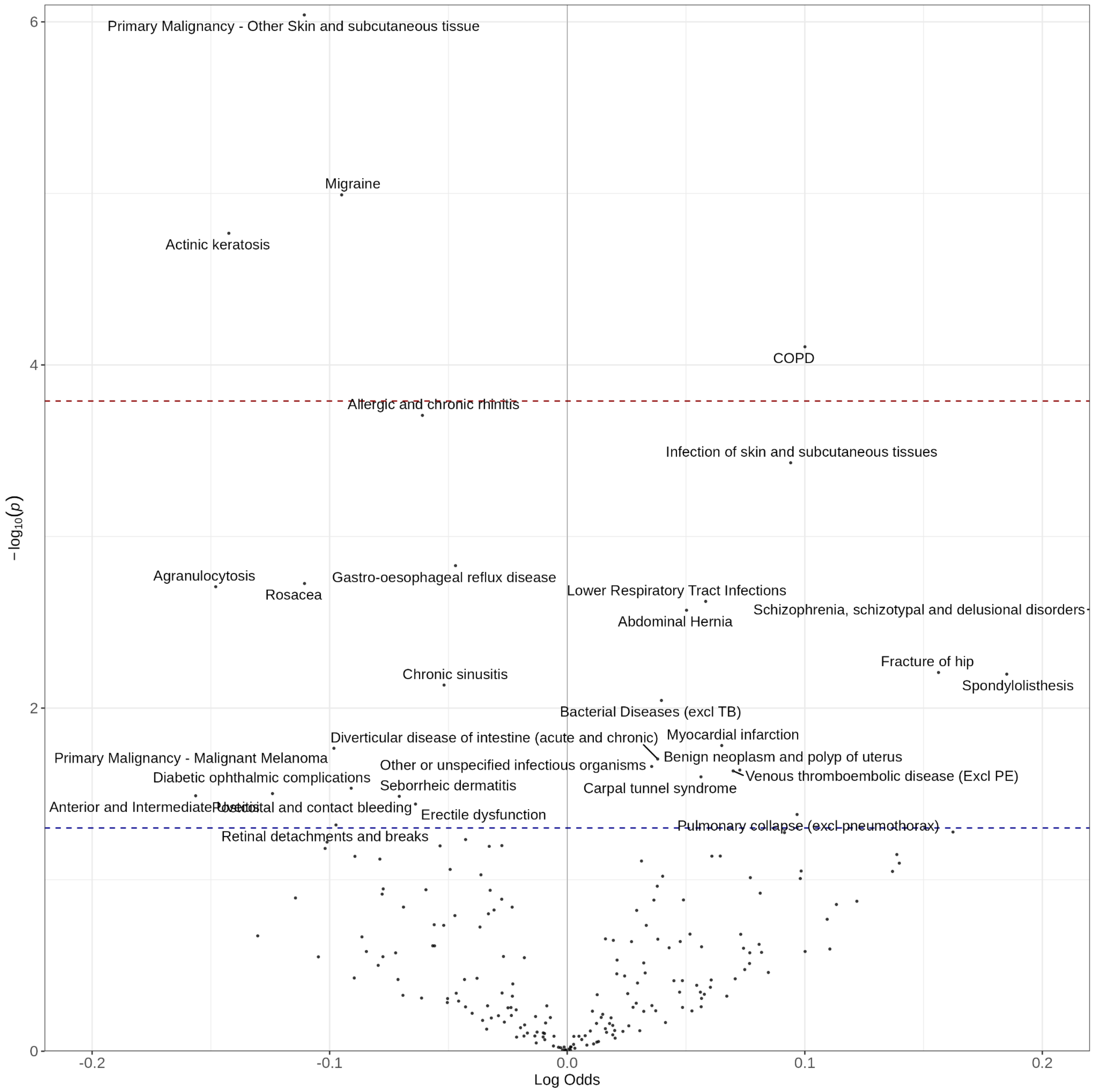


*Supplementary Figure 4. Volcano plot summarising phenome-wide association study results, showing potential associations between retinal pigment score (RPS) with 308 health conditions. The dashed blue line indicates p=0.05, while the dashed red line indicates the Bonferroni-adjusted significance threshold p=0.05/308 tests. Log odds are per standard deviation increase in RPS.*
